# The impact of increasing the minimum legal drinking age to 20 years in Lithuania on all-cause mortality – an interrupted time-series analysis

**DOI:** 10.1101/2021.04.07.21255080

**Authors:** Alexander Tran, Huan Jiang, Shannon Lange, Michael Livingston, Jakob Manthey, Maria Neufeld, Robin Room, Mindaugas Štelemėkas, Tadas Telksnys, Janina Petkevičienė, Ričardas Radišauskas, Jürgen Rehm

## Abstract

**Aims:** To determine the effect of an alcohol policy change, which increased the minimum legal drinking age (MLDA) from 18 years of age to 20 years of age on all-cause mortality rates in young adults in Lithuania.

**Methods:** An interrupted time series analysis was conducted on a dataset from 2001 to 2019 (*n* = 228 months). The model tested the effects of the MLDA on all-cause mortality rates (deaths per 100,000 individuals) in 3 age categories (15-17 years old, 18-19 years old, 20-22 years old). Additional models that included GDP as a covariate and taxation policy were tested as well.

**Results:** There was a significant effect of the MLDA on all-cause mortality rates in those 18-19 years old, when modelled alone. Additional analyses controlling for the mortality rate of other age groups showed similar findings. Inclusion of confounding factors (policies on alcohol taxation, GDP) eliminated the effects of MLDA.

**Conclusions:** Although there was a notable decline in all-cause mortality rates among young adults in Lithuania, a direct causal impact of MLDA on all-cause mortality rates in young adults was not definitively found.

**Short Summary:** We investigated the effect of an increase in minimum legal drinking age (MLDA) on all-cause mortality in young adults (aged 18-19). MLDA had a negative effect on all-cause mortality (even when controlling for mortality rates in other age groups), however when confounding factors were included, these effects were attenuated.

## Background

On January 1, 2018, Lithuania increased its minimum legal drinking age (MLDA) from 18 years of age to 20 years of age. This change was accompanied by increased enforcement by obligating all retailers to request for age verification if a customer appears to be younger than 25 (Miščikienė et al., 2020). At the same time, there were other alcohol control measures enacted that were intended to reduce availability and ban marketing (Miščikienė et al., 2020). With the goal of reducing alcohol consumption and attributable harm in Lithuania, which was among the highest globally in 2010 (Manthey et al., 2019), the enactment of these measures was part of a WHO initiative. This initiative was to implement all three of the World Health Organization’s (WHO) “best buys” for alcohol control policy, taxation increases, availability restrictions and ban of marketing (World Health Organization, 2017), within a relatively short period of time (Rehm et al., 2019). While the “best buys” were initially intended to specifically address non-communicable diseases only, these interventions have been shown to impact other disease outcomes as well (e.g., traffic injury; (Rehm et al., 2020)) and they reduce the overall burden of alcohol-attributable disease and mortality substantially (Chisholm et al., 2018, Štelemėkas et al., 2021 (in press)).

MLDA is the minimum age a person needs to be in order to be allowed to drink alcohol under law (World Health Organization, 2021). MLDA and its enforcement is an important part of the “best buys”, and has been established in all 53 countries in the European Region of the WHO, albeit with different age thresholds (see Supplementary Table S1; based on (Neufeld et al., 2021, World Health Organization, 2018)), between 16 and 21 years of age, and in some countries varying by beverage categories (beer, wine, spirits), or based on whether consumption is on- and off-premises. Moreover, some countries have separate age thresholds for purchasing vs. consuming alcohol. For most alcoholic beverage categories, the MLDA is 18 years, with the following mean levels: 18.0 years for beer and wine, and 18.2 years for spirits. After the recent change in 2018, Lithuania has one of the higher MLDAs in this region, with only Kazakhstan and Turkmenistan surpassing it with MLDAs of 21 years. The age restriction in Lithuania applies to the ability to purchase as well as to consume alcohol (Miščikienė et al., 2020).

MLDA has two main purposes: to reduce drinking and alcohol-attributable problems; alcohol-attributable disease and injury are major contributors to the latter. To date, two reviews have shown that MLDA generally achieves this goal, with most of the research based on the increase from 18 to 21 years in the United States of America (US), which became nationwide in 1984 (DeJong et al., 2014, Wagenaar et al., 2002). Enforcement was found to play a crucial role in the effectiveness of MLDA (Wagenaar et al., 2005).

In two of the most comprehensive recent studies on the effectiveness of MLDA, Carpenter and Dobkin (Carpenter et al., 2011, Carpenter et al., 2015) estimated the effects of increasing the US MLDA reduced night-time traffic fatalities among 18-20-year-olds by 17%, reduced suicide mortality and had substantial impacts on arrest rates across a wide range of crimes, including drink-driving, assault and robbery. These effects were likely mediated by reduced alcohol use, including but not limited to heavy episodic drinking. Studies from outside North America have corroborated the impact of MLDA in both directions; i.e., introductions and increases of MLDA were associated with reduced consumption and attributable harm, whereas lowering the MLDA (or minimum purchasing age) was associated with increased consumption, self-reported problems, traffic injuries and deaths, hospital admissions and rates of juvenile crime (Gruenewald et al., 2015, Huckle et al., 2014, Jiang et al., 2015).

The objective of the current study was to estimate the impact of the increase in MLDA on all-cause mortality in Lithuania among individuals 15-22 years of age, with the hypothesis that the increase in MLDA was associated with a significant decrease in mortality rates, particularly among those 18-19 years of age. We first tested this hypothesis using an interrupted time-series methodology (ITS; (Beard et al., 2019)), and then weo ran additional models that included a control series of younger (15-17 years) and older age groups (20-22 years), as well as an economic indicator. In addition, we conducted sensitivity analyses that tested the general trends in mortality through different time periods, as well as analyses focusing on men only.

## Methods

### Data

Monthly all-cause mortality data (monthly number of deaths) from 2001 to 2019 were obtained from Statistics Lithuania (Statistics Lithuania, 2021) by the Lithuanian University of Health Sciences (*n* = 228). The mortality data were separated into broad categories that represented disease and injury causes of death attributable to alcohol (Rehm et al., 2017): cancer, stroke, heart disease, other cardiovascular disease, diabetes and kidney diseases, gastrointestinal diseases, traffic injuries, unintentional injuries, intentional injuries and all other causes. The data were comprised of deaths for individuals between the ages of 15-22 years, which were separated into three age categories: 15-17 years of age, 18-19 years of age, and 20-22 years of age, as well as by sex. Injuries are the main cause of deaths in these age-groups (see Figure 1).

**Figure 1:**
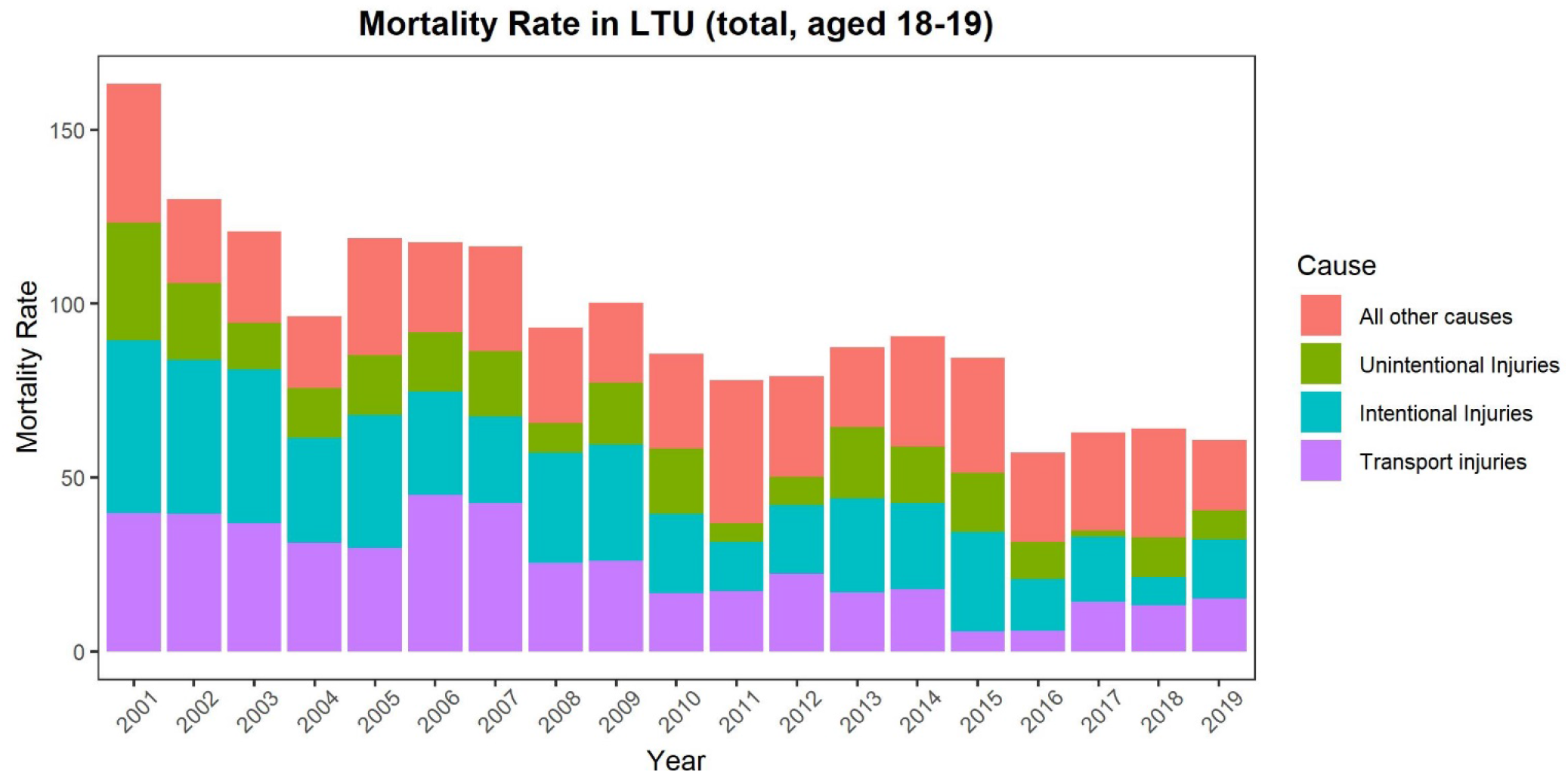
Mortality rate in Lithuania in the age group 18- and 19-year-olds, 2000-2019 separated by 4 major death categories

> Yearly population data were also obtained from Statistics Lithuania for the individuals 15 to 22 years of age and were combined into the three age categories described above (aged 15-17, aged 18-19, and aged 20-22). These yearly data were linearly imputed to obtain monthly population data used to calculate the rates (see below). To control for potential economic confounds, quarterly data on gross domestic product (GDP) *per capita* (in USD) were obtained from Statistics Lithuania, and converted into monthly gross domestic product also using linear imputation (Lithuanian Department of Statistics, 2021).

### Dependent Variable

Crude mortality rates (monthly deaths per 100,000 individuals) were calculated for each of the three age groups, by sex and for both sexes combined. As the age-categories were small, no age-adjustment was applied. For the direct control we used the difference between the mortality rates of the group of interest (aged 18-19) and of control groups (aged 15-17 or aged 20-22). We also performed an analysis for men (aged 18-19), controlling for women (aged 18-19) by creating a difference score between the mortality rates.

### Predictor variable and covariate

The primary variable to be tested was the above-described increase in MLDA on January 1, 2018. This policy change was part of an implementation of three alcohol control measures at the same time (ban on advertising, reduced off-premises sales hours and MLDA; (Miščikienė et al., 2020)) which, as mentioned above, was part of an overall strategy of implementing all WHO “best buys” for alcohol control policy within a short period of time (Rehm et al., 2019). Thus, different models were estimated:

- An ITS analysis of mortality rates among the total population, aged 18-19 with only the 2018 policy implementations;
- ITS analyses estimating the effect of the policy on mortality rates of the total population, aged 18-19 by directly controlling for the effects of those aged 15-17, and those aged 20-22, by creating new dependent variables of the difference between the respective time series of mortality rates; similarly, we used the time series of mortality rates of women (aged 18-19) as a control for mortality rates of men (aged 18-19);
- An ITS analysis additionally controlling for the immediate effects of the most powerful policy intervention on all-cause mortality rates: a substantial increase in excise taxation of 111-112% for wines and beer and 23% for ethyl alcohol, mainly applying to spirits implemented in March 2017 (Rehm et al., 2021, Štelemėkas et al., 2021 (in press)) and for economic confounds (GDP per capita)
- An ITS analysis of four different time periods with and without major alcohol policy measures, defined as follows: (Period 1) January 2001 until December 2007 (no major alcohol control policies introduced; several small policies associated with increased availability); (Period 2) January 2008 until December 2009 (year of sobriety; five alcohol control policies introduced); (Period 3) January 2010 until March 2014 (no major alcohol control policies introduced); and (Period 4) April 2014 until December, 2019 (total of nine alcohol control policies introduced; see (Miščikienė et al., 2020, Rehm et al., 2021).

Alternatively, we did a sensitivity analyses using the following periods which included only major policies as breakpoints, and thus did not consider the policies of 2014 to 2016 as periods of high intensity of alcohol control policy: (Period 1) January 2001 until December 2007; (Period 2) January 2008 until December 2009; (Period 3) January 2010 until February 2017; and (Period 4) March 2017 until December 2019

In addition, we calculated models controlled by GDP *per capita* and for men only (women only models were not possible due to small numbers). Men made up 79.27% of deaths in the age group of 18-to 19-year-olds, averaged over the time period.

All alcohol control measures used in our models were dummy coded variables with a value of 0 before the measure, and 1 thereafter, thus testing for an abrupt permanent effect.

### Statistical analyses

An interrupted time series analysis was then employed using a general additive mixed model (GAMM; (Beard et al., 2019) to test the effect of increased MLDA (January 2018). In this model, seasonality was controlled for using a single smoothing term that was a 12-knot cubic spline (monthly seasonal trend). The model residuals were checked for normality and stationarity using QQ plots and the augmented Dickey-Fuller (ADF) test. To achieve stationarity, and to remove autocorrelation in the time series, an autoregressive and integrated moving average (ARIMA) model for errors was used when necessary (details on all the models can be found in the supplementary materials in Supplementary Table S2 to S5).

All analyses were performed using R version 4.0.4 (R Project, 2021).

## Results

Once the data were fit with AR and MA terms, they approximated normality and stationarity (see Supplemental Figure S4). Overall, there appeared to be a general decline in all-cause mortality rate across all three age groups since January 2001. Figure 1 gives an overview of the time trend in all-cause mortality rate among those aged 18-19, separated and grouped by major causes of death. Given that various causes of death had very small contributions to all-cause mortality, only 4 categories were selected: unintentional injuries, intentional injuries, transport injuries, and all other causes of death (Supplemental Figure S1 in the Supplementary Materials shows the mortality rates, separated by all 9 categories, as well as Supplemental Figure S2 and S3 in the Supplementary Materials, which shows the 4-category breakdown for the two other age groups). As demonstrated in Figure 1, injury was responsible for the majority (70.01%) of deaths of deaths in those aged 18-19. It should be noted that deaths by suicide constitute to a large portion of the intentional injury deaths in Lithuania. For instance, Lithuania’s age-standardize suicide rate is 25.7 per 100 000 and therefore twice as high as the rate of the WHO European Region, which is 12.9 per 100 000 (World Health Organization, 2019).

### Interrupted ITS models with mortality rates as dependent variable

In the most parsimonious model (testing the effect of the increased MLDA in 2018 on mortality rates alone) there was a marginal, non-significant effect in the 15-17 age group (t(226) = -1.76, p = .079), a significant effect in the 18-19 age group (t(226) = -3.05, p = .0025), and a significant effect in the 20-22 age group (t(226) = -3.65, p = .00035; details on models in the Supplementary Materials). However, when GDP *per capita* was introduced, this variable became significant and the effect for the MDLA policy enactment of January 2018 disappeared (see Supplementary Table A2). As men make up the majority of deaths in this age group, their ITS models showed very similar results (see Supplementary Table A2).

When adding the taxation policy in 2017 to the model, there was no effect of the MLDA policy in 2018 on mortality rate, either in those aged 18-19 or in the other two age groups (see Appendix Table A3). But there was a marginal effect of the taxation policy in 2017 for those aged 15-17 (t(224) = -1.90, p = .059), and a significant effect for those aged 20-22 (t(224) = -2.24, p = .026).

For all ITS models, GDP *per capita* had a significant negative effect on mortality rates, such that as GDP increased, mortality rates decreased. The sign of this effect was different from the model for all age groups (Štelemėkas et al., 2021 (in press)). Overall, even for the models that included all variables considered, there was a relatively poor fit for the 15-17 age group (R^2^-adjusted = 0.22) as well as the 18-19 age group (R^2^-adjusted = 0.27), while the 20-22 age group had a better fit (R^2^-adjusted = 0.41).

### Interrupted time series models with direct control by other time series

Table 1 gives an overview of all models, which used the time series of other age groups and genders as direct controls via differencing.

**Table 1:**
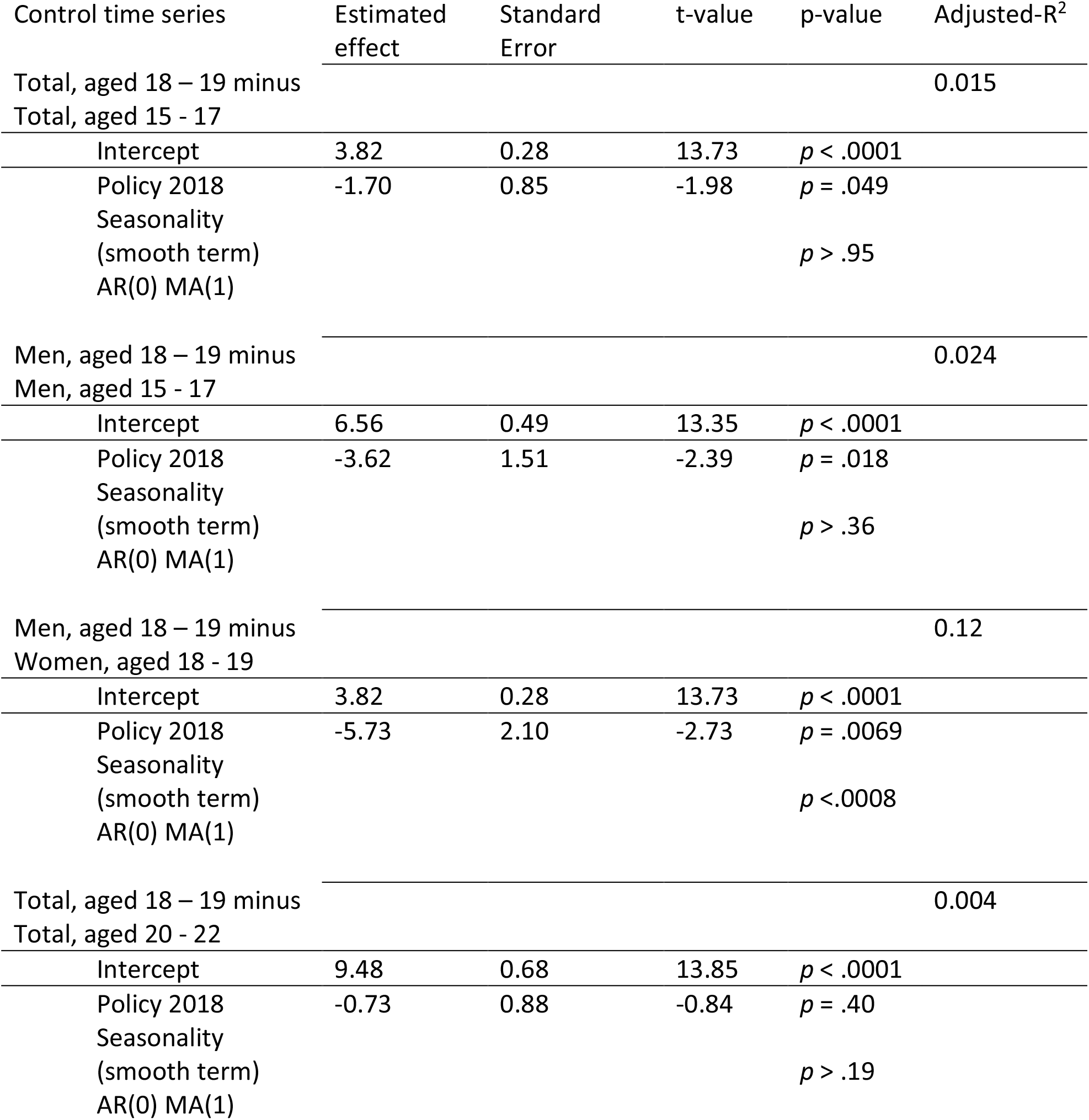
Overview of ITS of MLDA (January 2018) on mortality rates controlling for time series of other populations

| Control time series | Estimated effect | Standard Error | t-value | p-value | Adjusted-R<sup>2</sup> |
| --- | --- | --- | --- | --- | --- |
| Total, aged 18 – 19 minus<br>Total, aged 15 - 17 |  |  |  |  | 0.015 |
| Intercept | 3.82 | 0.28 | 13.73 | $p < .0001$ | |
| Policy 2018 | -1.70 | 0.85 | -1.98 | $p = .049$ | |
| Seasonality<br>(smooth term) | | | | $p > .95$ | |
| AR(0) MA(1) |  |  |  |  |  |
| Men, aged 18 – 19 minus<br>Men, aged 15 - 17 |  |  |  |  | 0.024 |
| Intercept | 6.56 | 0.49 | 13.35 | $p < .0001$ | |
| Policy 2018 | -3.62 | 1.51 | -2.39 | $p = .018$ | |
| Seasonality<br>(smooth term) | | | | $p > .36$ | |
| AR(0) MA(1) |  |  |  |  |  |
| Men, aged 18 – 19 minus<br>Women, aged 18 - 19 |  |  |  |  | 0.12 |
| Intercept | 3.82 | 0.28 | 13.73 | $p < .0001$ | |
| Policy 2018 | -5.73 | 2.10 | -2.73 | $p = .0069$ | |
| Seasonality<br>(smooth term) | | | | $p < .0008$ | |
| AR(0) MA(1) |  |  |  |  |  |
| Total, aged 18 – 19 minus<br>Total, aged 20 - 22 |  |  |  |  | 0.004 |
| Intercept | 9.48 | 0.68 | 13.85 | $p < .0001$ | |
| Policy 2018 | -0.73 | 0.88 | -0.84 | $p = .40$ | |
| Seasonality<br>(smooth term) | | | | $p > .19$ | |
| AR(0) MA(1) |  |  |  |  |  |

**Table 2:**
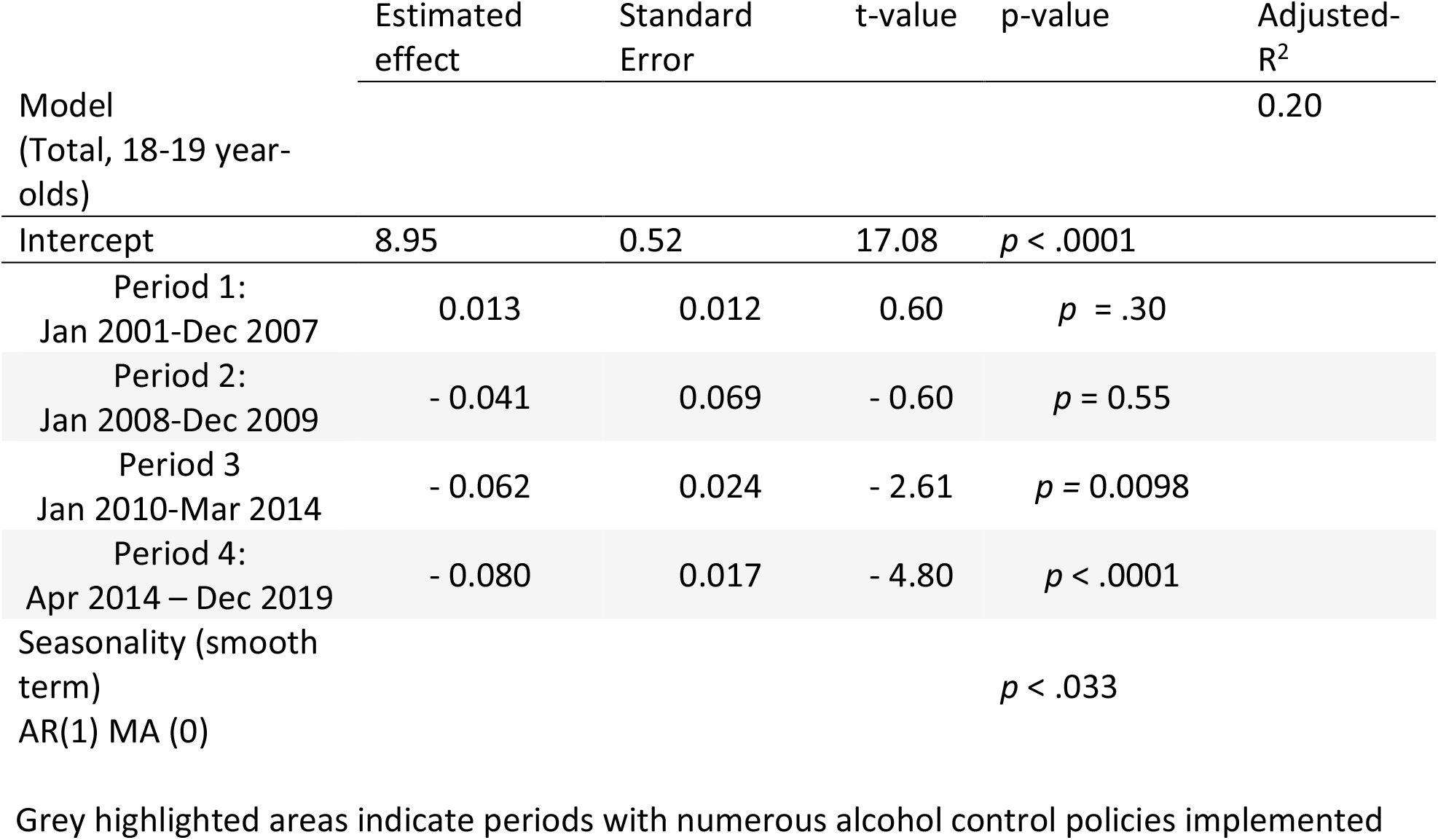
Estimated trends in mortality rates during four different periods of alcohol control policy in Lithuania

The effect of the 2018 MLDA policy persisted after controlling for the mortality rate of those aged 15-17, and when taking the mortality rate in women as a control for the mortality rate in men. However, when controlling for those aged 20-22, the effect was no longer significant.

### Comparing periods with different intensity of alcohol policy enactments

Periods during which alcohol control policies were implemented (i.e., Period 2: January 2008 – December 2009, and Period 4: April 2014 – December 2019) were associated with trend changes in mortality rates among 18-to 19-year-olds. In the first such period, Period 2: January 2008 – December 2009, the trend changed from a slightly positive slope in mortality rates into a negative slope (i.e., from slightly increasing to decreasing). Furthermore, in the most recent period, Period 4: April 2014 - December 2019, there was a highly significant decrease in all-cause mortality rate.

The sensitivity analysis corroborated these results, again, the wider defined periods with alcohol policy implementation were associated with change of trends and the most pronounced declining mortality trend, respectively (details see Appendix Table A2). And not surprisingly, the same pattern of results was found to be even more pronounced in men (for details see Appendix Table A3).

## Discussion

Overall, there was a decreasing trend in the all-cause mortality rates among individuals aged 18-19 in Lithuania, the level of which was associated with implementation of alcohol control policies. The exact role that the increase in MLDA played in this trend remains unclear, as a number of other alcohol control policies were implemented in close temporal proximity, as well there was a possible confounding effect of GDP. While an ITS without additional confounders, and the controlled ITS (age-differenced time series) showed a significant effect of MLDA, models with additional confounders were no longer significant.

The following potential limitations should be considered: First, Lithuania is a small country, and the mortality numbers for the age groups likely to be impacted by increasing the MLDA, such as 18-to 19-year-olds, are relatively small. Furthermore, there was a change in population registration procedures resulting in a decline in the recorded population in Lithuania over the observed period. Consequently, fluctuations of only a few deaths per month made a large difference in mortality rates (deaths per 100,000) and may explain the relatively poor model fit. The small number of deaths per month, with several months of zero deaths, were also the reason we could not separate models for women. Second, alcohol policy implementation activity happened in clusters of periods (see methods for detailed explanation), so within the 20-year window we could distinguish four different periods, two with low or fewer alcohol control policies and two with stricter alcohol policy implementation. To separate out the effects of single policies such as MLDA is hard, especially, when the increase in MLDA was coupled with a ban on advertising and an availability restriction. While the ban on advertising may not have impacted the mortality rate abruptly in the short term (Babor et al., 2010), it still may have contributed to the results. Furthermore, as the alcohol policy implementation happened in close temporal proximity to the 2017 policy change, the overlap was large (Pearson r = 0.81), creating potential multicollinearity. Finally, although we attempted to remove confounds by controlling for those aged 15-17 and those aged 20-22 there may have been some issues with these models. On the one hand, young people intermingle, and it can very well be that they go out in groups together and would collectively be impacted by certain behaviors such as drink-driving (Thombs et al., 1997, Brown, 1998, González-Iglesias et al., 2015). Furthermore, it has been shown in other countries that simply the announcement of shifting MLDA may sensitize teenagers/young adults and their families to the issue of alcohol and attributable harm, thus widening the impact of the law to neighboring age groups or shifting the effects to some months prior to actual implementation. For instance, Møller showed that the introduction of legal restrictions, i.e., the implementation of a MLDA in Denmark, resulted in a reduction of alcohol use not only in the age group targeted but also among older kids (Møller, 2002).

These limitations make it hard to unequivocally attribute a decrease on mortality among 18-to 19-year-olds to the increase of a MLDA. However, it is possible to indicate that the various alcohol control policies introduced in a period since March 2014 to December 2019 were strongly associated with this reduction of total mortality in this age group. Given the leading causes of death in this age-group (unintentional injuries), these national policies were aimed at reducing drink driving, such as the ban on alcohol sales at petrol stations in 2016, zero tolerance for alcohol use among newly-licensed drivers, or making a blood alcohol concentration above 1.5 ‰a criminal offense (for a detailed description of all policies see (Miščikienė et al., 2020). In addition, it has been shown that universal policies such as taxation increases or reductions of availability also reduce traffic injury and fatalities (Rehm et al., 2020). These types of universal policies, and policies targetting drink-drinking behaviour would have impacted mortality rates in those 18-to 19 years old by reducing consumption and binge drinking, and high-risk behaviours respectively, both of which are strongly related to all kinds of injury (Cherpitel et al., 2019, Rehm et al., 2003).

In conclusion, while we could not specifically establish a causal impact of the increase of MLDA in Lithuania on total mortality, overall the recent implementation of various alcohol control policies was associated with a decrease in mortality over and above secular trends.

## Data Availability

The R Code used to analyze and compute variables for the current study are available from the corresponding author upon reasonable request. The data may be obtained on request through the Lithuanian Governmental institutions (Lithuanian Institute of Hygiene, Statistics Lithuania).

## Acknowledgments

Research reported in this publication was also supported by the National Institute on Alcohol Abuse and Alcoholism of the National Institutes of Health (NIAAA) [Award Number 1R01AA028224-01]. JR acknowledges funding from the Canadian Institutes of Health Research, Institute of Neurosciences, and Mental Health and Addiction (CRISM Ontario Node grant no. SMN-13950).

## Conflict of Interest Statement

The Authors declare that there are no conflicts of interest. The authors alone are responsible for the views expressed in this publication and they do not necessarily represent the decisions or the stated policy of the World Health Organization, of which Maria Neufeld is a consultant

## Data Availability Statement

The R Code used to analyze and compute variables for the current study are available from the corresponding author upon request. The data may be obtained on request through the Lithuanian Governmental institutions (Lithuanian Institute of Hygiene, Statistics Lithuania).

## Notes

### Competing Interest Statement

The authors have declared no competing interest.

### Funding Statement

Research reported in this publication was supported by the National Institute on Alcohol Abuse and Alcoholism of the National Institutes of Health (NIAAA), grant number 1R01AA028224-01. This research was conducted as part of the project "Evaluation of the impact of alcohol control policies on morbidity and mortality in Lithuania and other Baltic states" and we would like to thank the whole team for their input to wider discussions in generating the research reported in this paper. Content is the responsibility of the authors and does not reflect official positions of NIAAA or the National Institutes of Health.

### Author Declarations

This study was approved by the CAMH Research Ethics Board (REB).

