## Supplemental Materials for "The impact of increasing the minimum legal drinking age to 20 years in Lithuania on all-cause mortality – an interrupted time-series analysis"

### Supplementary Figure S1: Number of deaths in Lithuania in the age group 18-to-19 year-olds, 2001-2019 by all 9 death categories

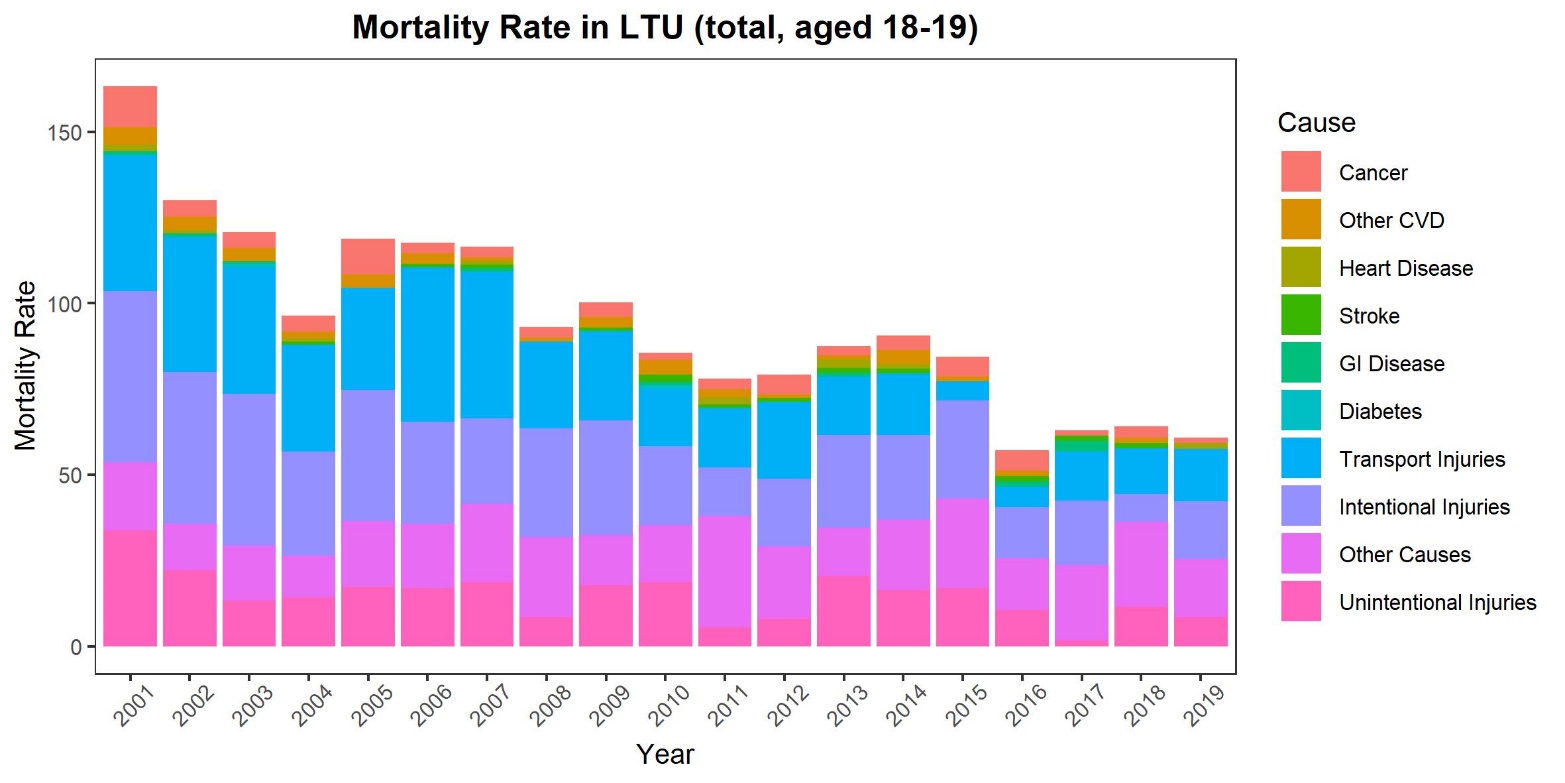

### Supplementary Figure S2: Number of deaths in Lithuania in the age group 15- to 17-year-olds, 2000-2019 by 4-major death categories

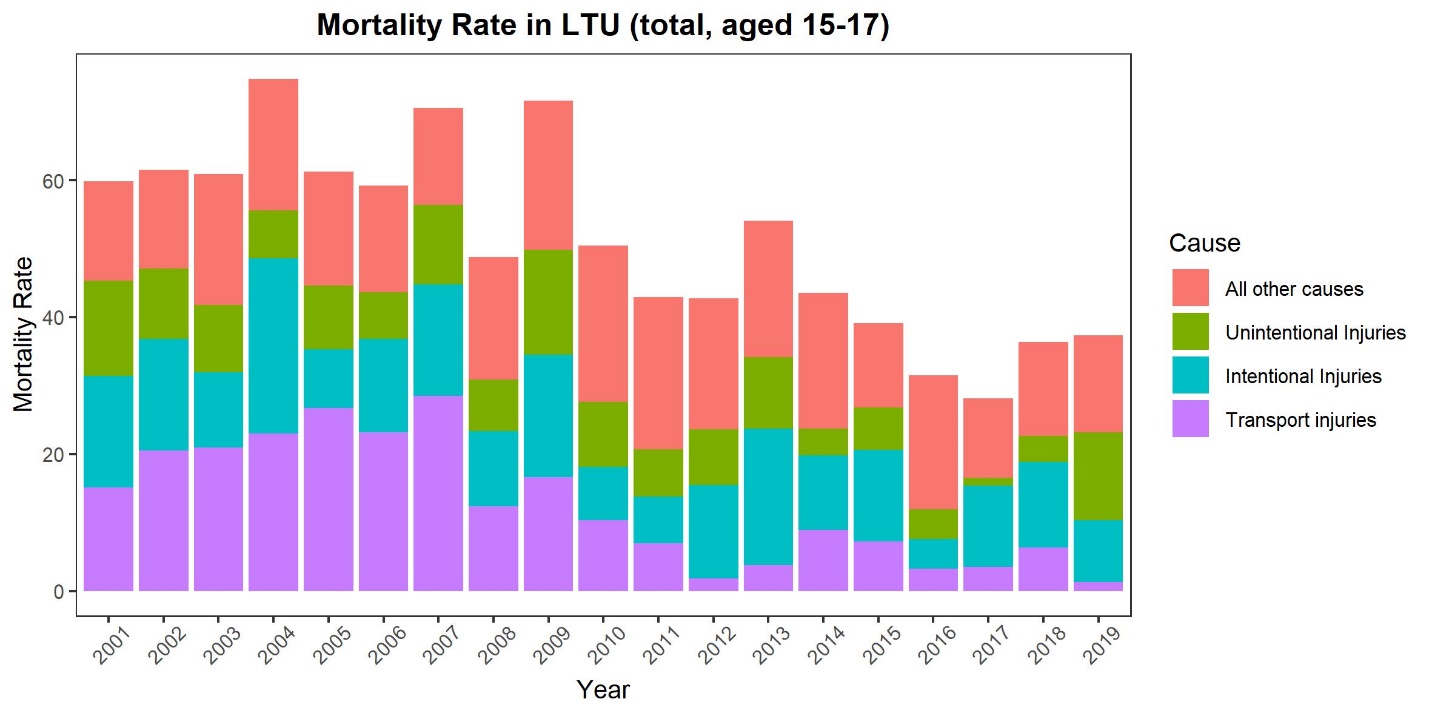

### Supplementary Figure S3: Number of deaths in Lithuania in the age group 20- to 22-year-olds, 2000-2019 by 4-major death categories

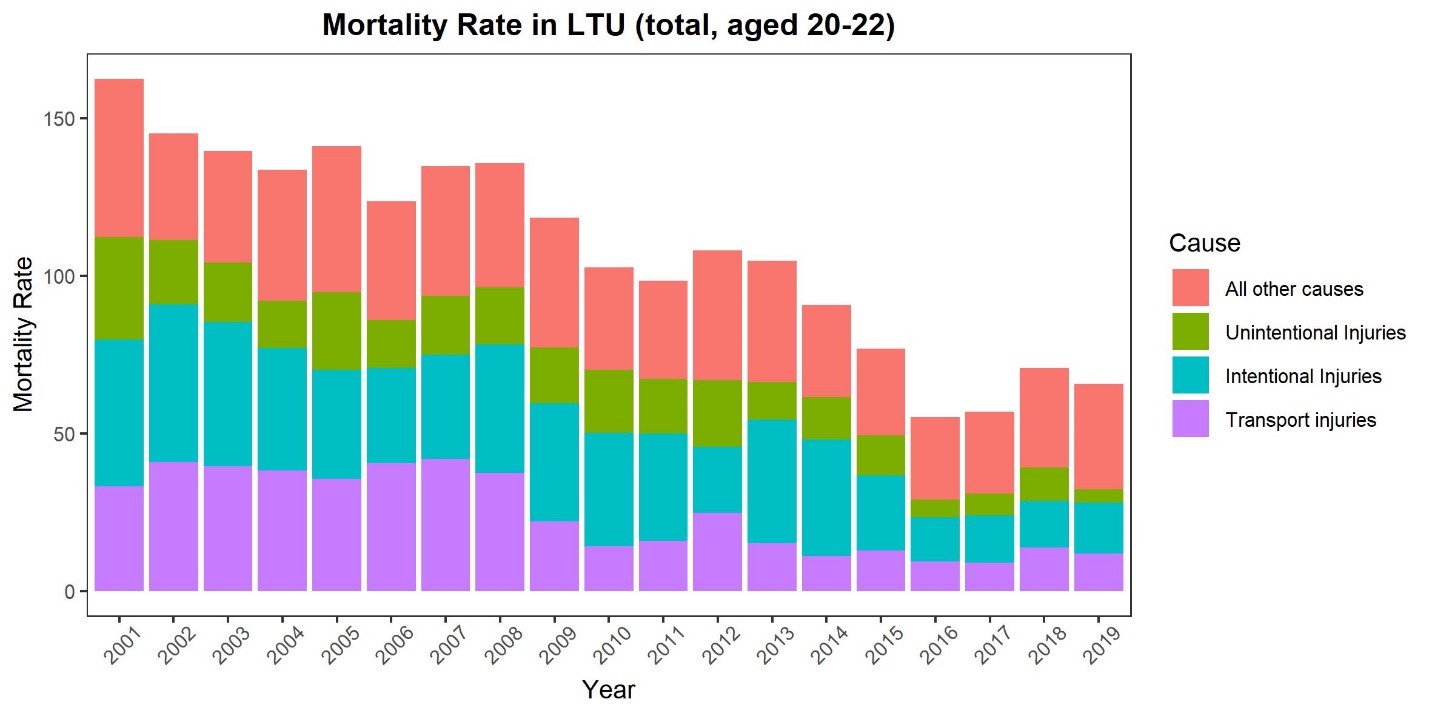

### Supplementary Figure S4: Check for normality and autocorrelation of residuals

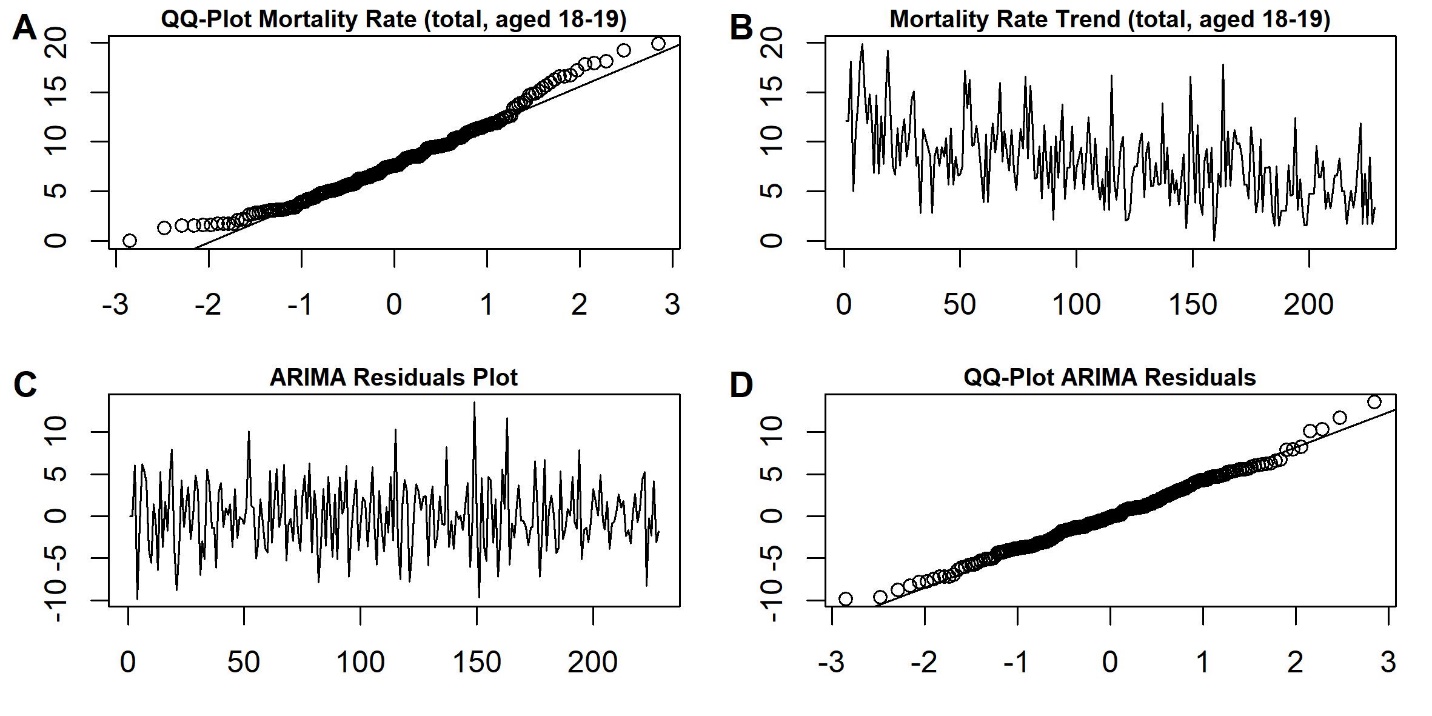

A: QQ-Plot for mortality rates in total population, aged 18-19, B: Time series trend of mortality rate (stationary based on ADF-test = -6.03, *p* < .01), C: Residual plot from ARIMA model (1,1,0), D: QQ-plot for ARIMA residuals.

### SupplementaryFigure S5: ITS Models of mortality rate in age group 15- to 17-year olds, Model 1: MLDA only, Model 2: Taxation policy (2017), MLDA policy (2018), and GDP

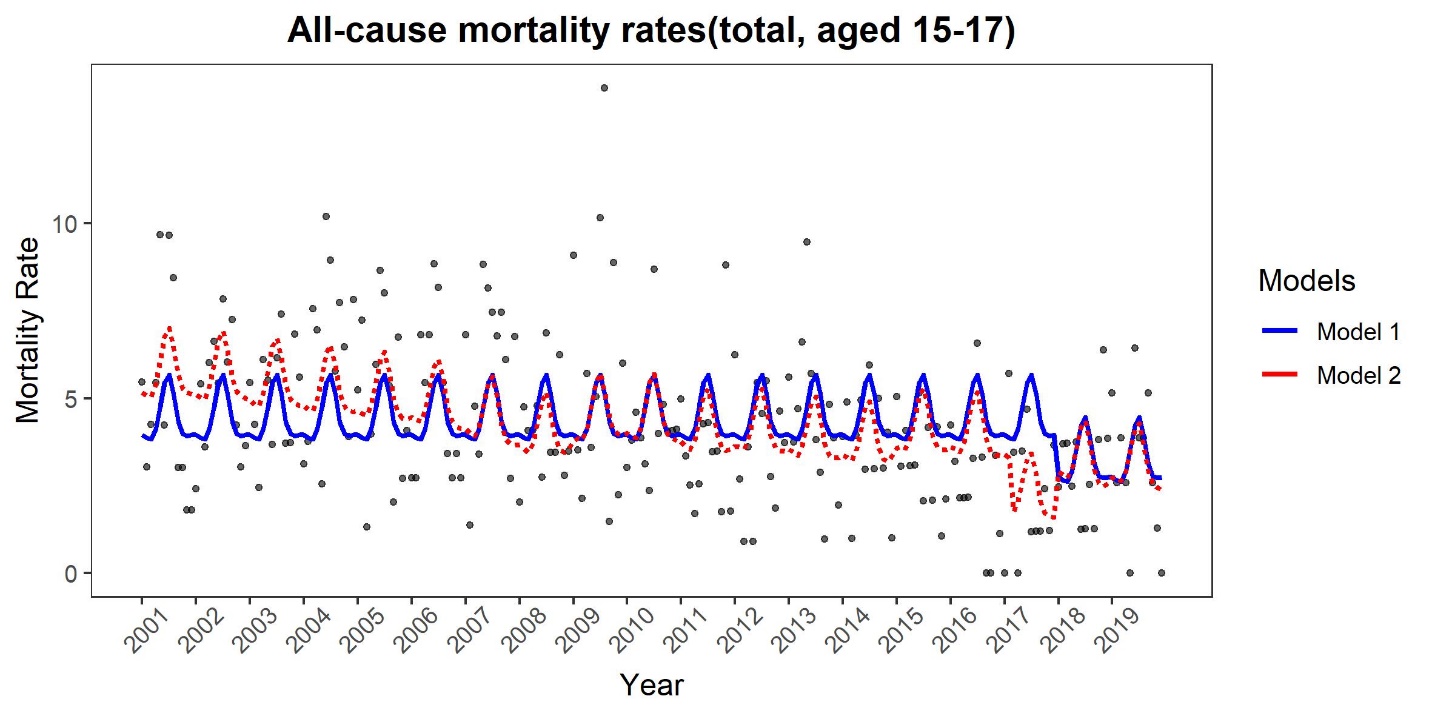

Model estimates compared to mortality data in those aged 15 – 17, where Model 1 includes MLDA, seasonality and autocorrelation, and Model 2 includes taxation, MLDA, GDP, seasonality and autocorrelation. See Supplementary Table S4 for full model details.

### Supplementary Figure S6: ITS Models of mortality rate in age group 18- to 19-year olds, Model 1: MLDA only, Model 2: Taxation policy (2017), MLDA policy (2018), and GDP

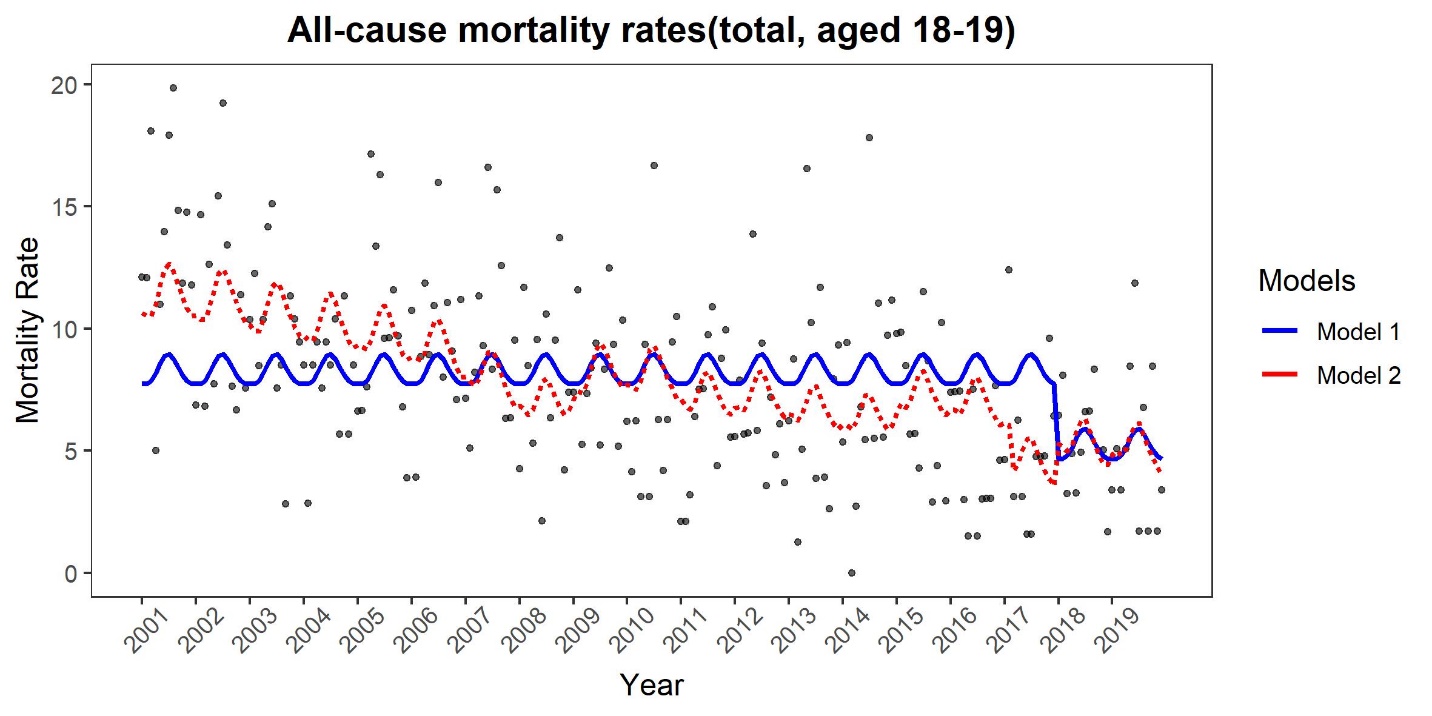

Model estimates compared to mortality data in those aged 18 – 19, where Model 1 includes MLDA, seasonality and autocorrelation, and Model 2 includes taxation, MLDA, GDP, seasonality and autocorrelation. See Supplementary Table S3 for full model details.

### Supplementary Figure S7: ITS Models of mortality rate in age group 20- to 22-year olds, Model 1: MLDA only, Model 2: Taxation policy (2017), MLDA policy (2018), and GDP

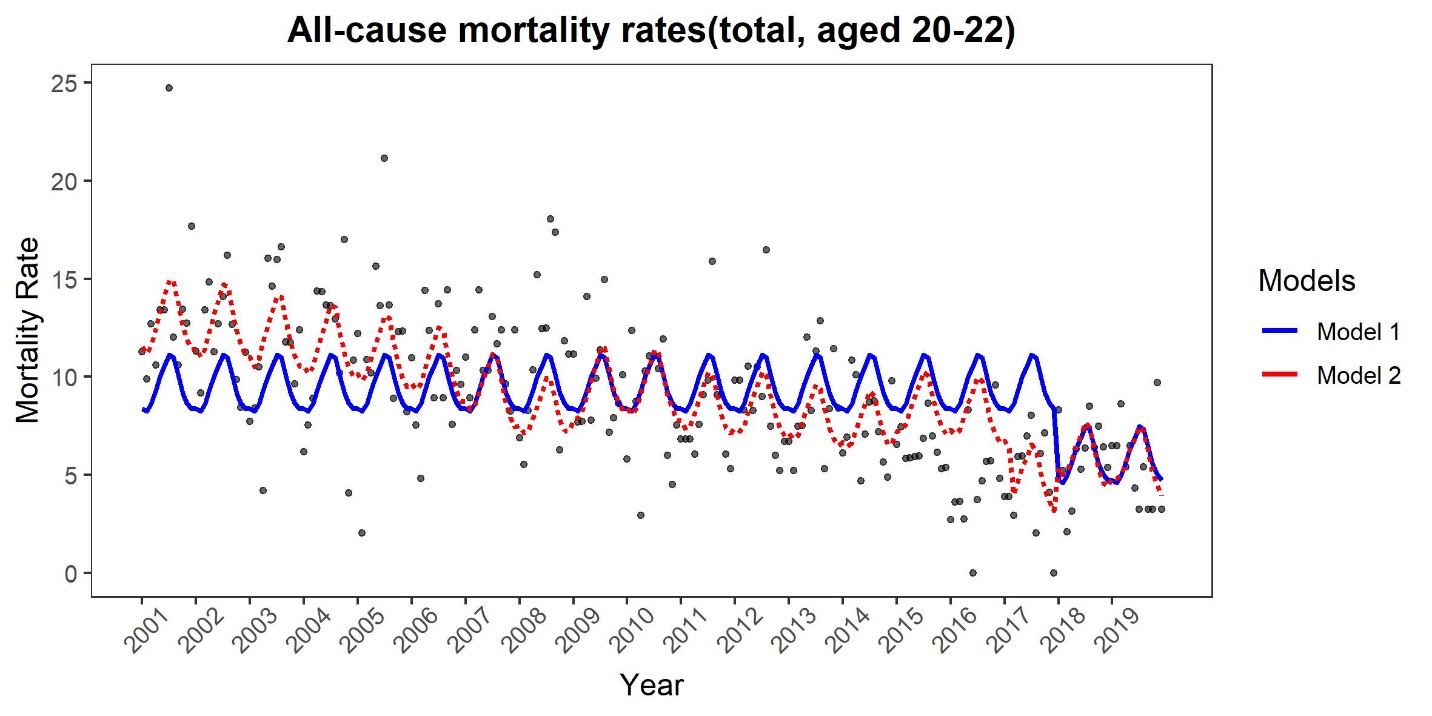

Model estimates compared to mortality data in those aged 20 – 22, where Model 1 includes MLDA, seasonality and autocorrelation, and Model 2 includes taxation, MLDA, GDP, seasonality and autocorrelation. See Supplementary Table S5 for full model details.

### Supplementary Figure S8: Differenced time series in Table 1

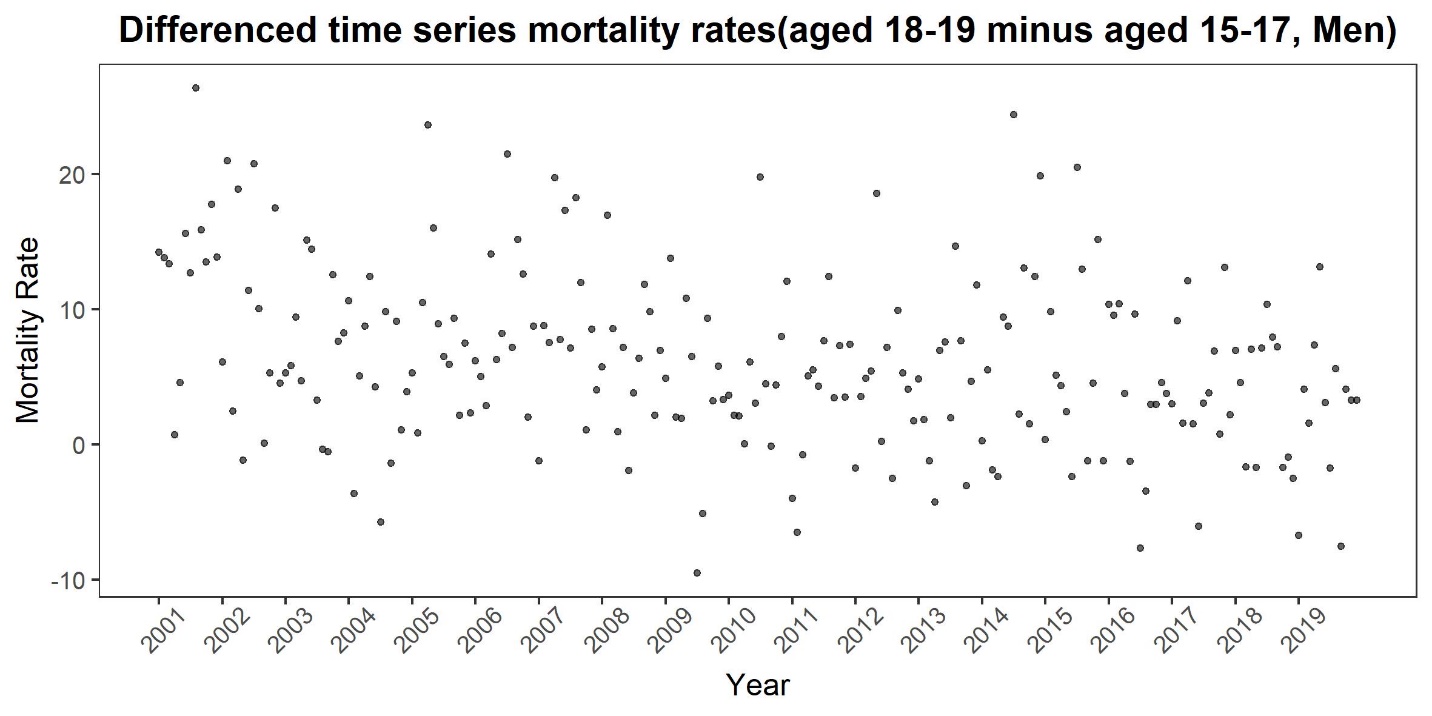

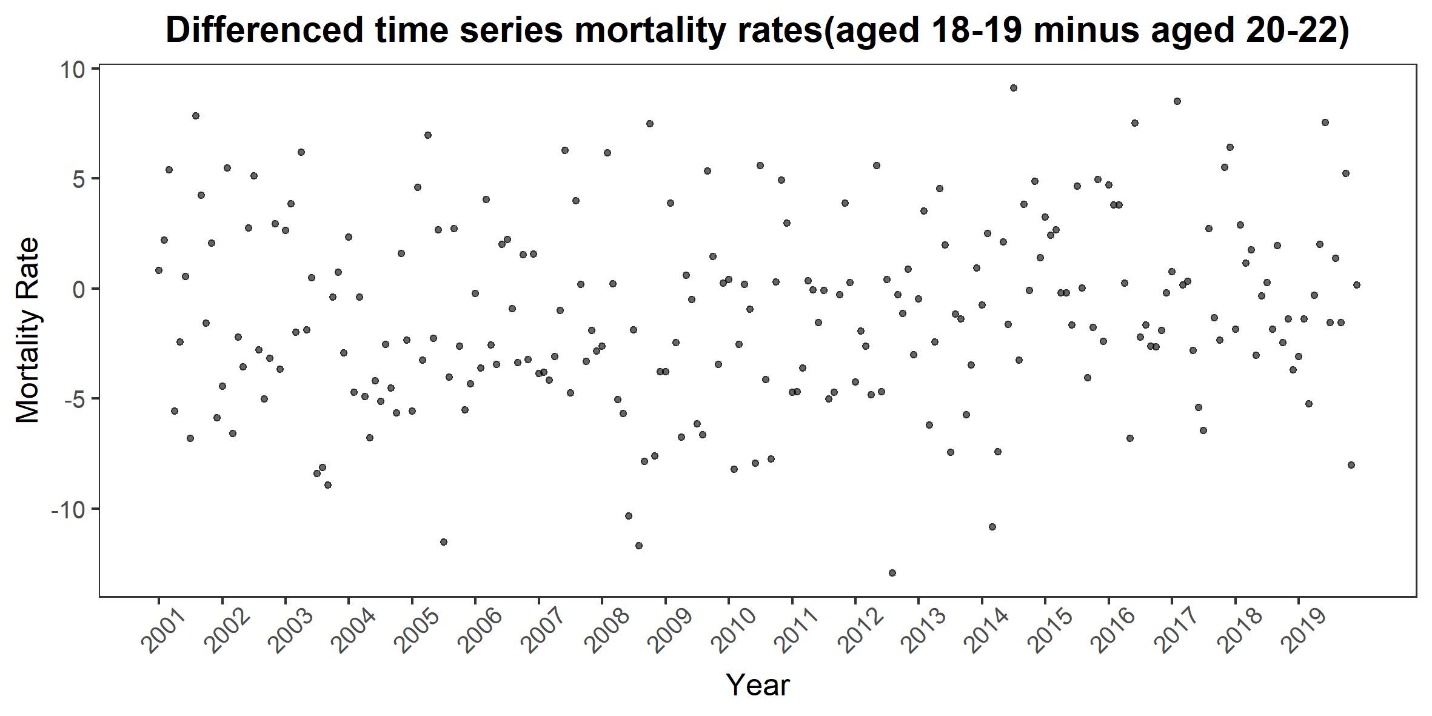

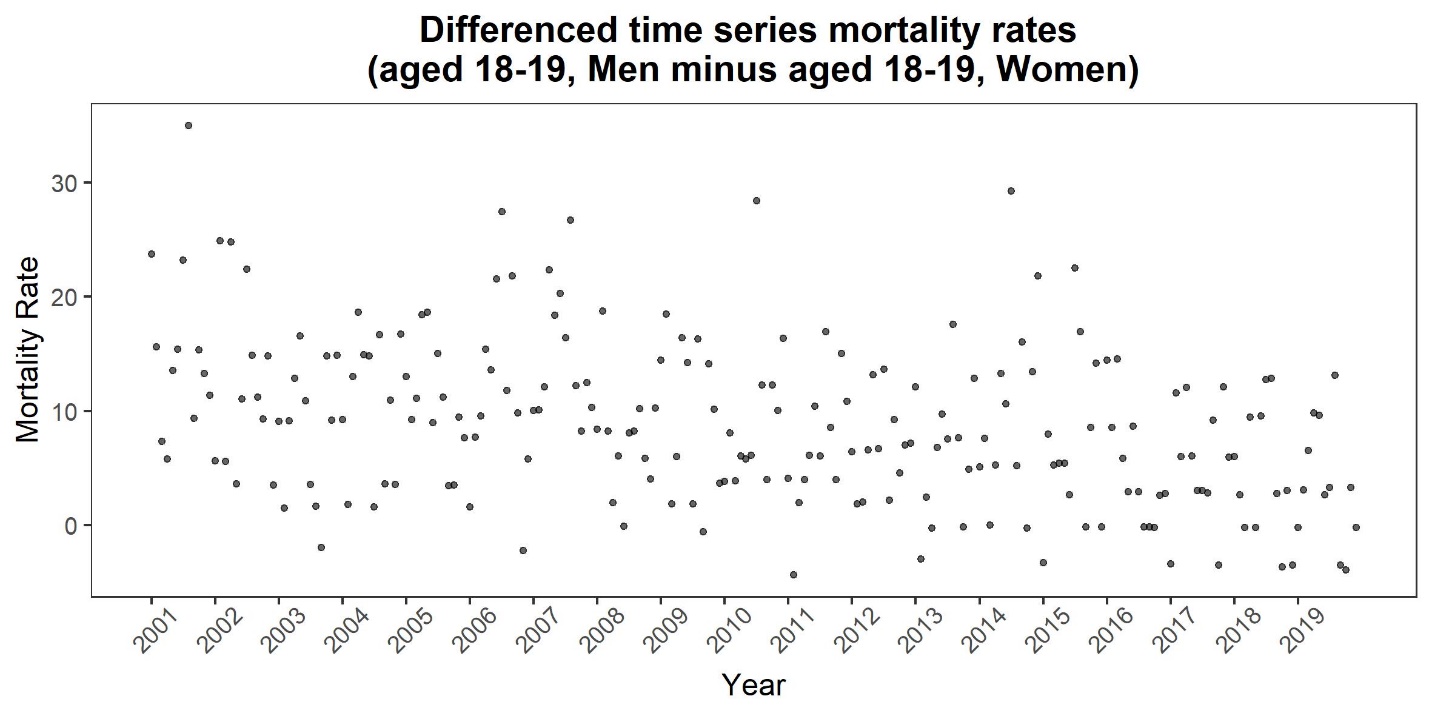

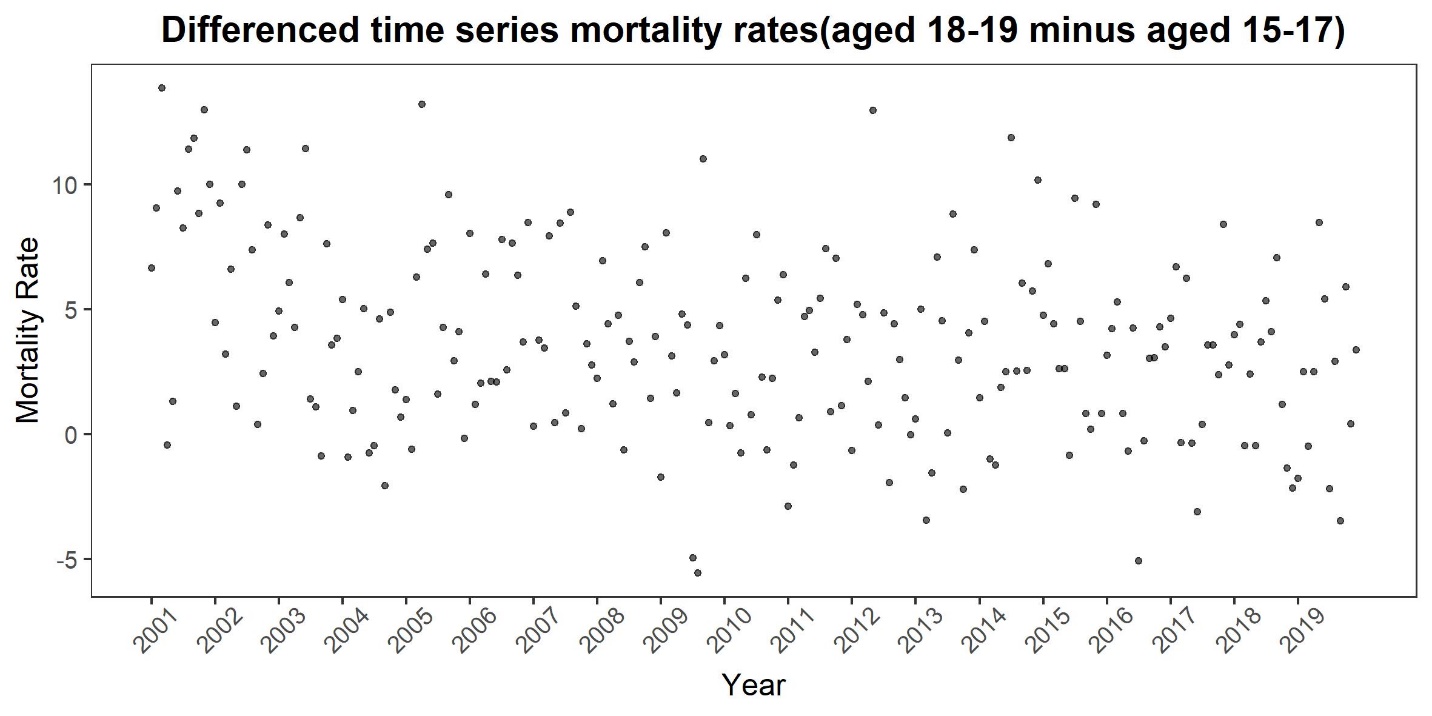

### Supplementary Table S1: Beverage-specific MLDAs in countries of the WHO European Region

| **Country** | **Beer** | **Wine** | **Spirits** |
| --- | --- | --- | --- |
| Albania | 18 | 18 | 18 |
| Andorra | 18 | 18 | 18 |
| Armenia | 18 | 18 | 18 |
| Austria | 16 | 16 | 18 |
| Azerbaijan | 18 | 18 | 18 |
| Belarus | 18 | 18 | 18 |
| Belgium | 16 | 16 | 18 |
| Bosnia and Herzegovina | 18 | 18 | 18 |
| Bulgaria | 18 | 18 | 18 |
| Croatia | 18 | 18 | 18 |
| Cyprus | 17 | 17 | 17 |
| Czech Republic | 18 | 18 | 18 |
| Denmark | 16 | 16 | 18 |
| Estonia | 18 | 18 | 18 |
| Finland | 18 | 18 | 20 |
| France | 18 | 18 | 18 |
| Georgia | 18 | 18 | 18 |
| Germany | 16 | 16 | 18 |
| Greece | 18 | 18 | 18 |
| Hungary | 18 | 18 | 18 |
| Iceland | 20 | 20 | 20 |
| Ireland | 18 | 18 | 18 |
| Israel | 18 | 18 | 18 |
| Italy | 18 | 18 | 18 |
| Kazakhstan | 21 | 21 | 21 |
| Kyrgyzstan | 18 | 18 | 18 |
| Latvia | 18 | 18 | 18 |
| Lithuania | 20 | 20 | 20 |
| Luxembourg | 16 | 16 | 16 |
| Malta | 17 | 17 | 17 |
| Monaco | 18 | 18 | 18 |
| Montenegro | 18 | 18 | 18 |
| Netherlands | 18 | 18 | 18 |
| North Macedonia | 18 | 18 | 18 |
| Norway | 18 | 18 | 20 |
| Poland | 18 | 18 | 18 |
| Portugal | 18 | 18 | 18 |
| Republic of Moldova | 18 | 18 | 18 |
| Romania | 18 | 18 | 18 |
| Russian Federation | 18 | 18 | 18 |
| San Marino | 16 | 16 | 16 |
| Serbia | 18 | 18 | 18 |
| Slovakia | 18 | 18 | 18 |
| Slovenia | 18 | 18 | 18 |
| Spain | 18 | 18 | 18 |
| Sweden | 20 | 20 | 20 |
| Switzerland | 16 | 16 | 18 |
| Tajikistan | 18 | 18 | 18 |
| Turkey | 18 | 18 | 18 |
| Turkmenistan | 21 | 21 | 21 |
| Ukraine | 18 | 18 | 18 |
| United Kingdom | 18 | 18 | 18 |
| Uzbekistan | 20 | 20 | 20 |

### Supplementary Table S2: Effect of MLDA, taxation, and economic wealth (GDP) on mortality rates among all 18- to 19-year-olds in Lithuania

|  | Estimated effect | Standard Error | t-value | p-value | Adjusted-R^2^ |
| --- | --- | --- | --- | --- | --- |
| Model (Total) |  |  |  |  | 0.27 |
| Intercept | 12.97 | 0.69 | 18.80 | *p* < .0001 |  |
| GDP *per capita* | -0.0016 | 0.00022 | -7.32 | *p* <0001 |  |
| Policy 2018 | 0.20 | 0.87 | 0.23 | *p* = .82 |  |
| Seasonality (smooth term) |  |  |  | *p* < .0007 |  |
| AR(1) MA(0) |  |  |  |  |  |

| Model 2 (Total) |  |  |  |  | 0.27 |
| --- | --- | --- | --- | --- | --- |
| Intercept | 12.76 | .70 | 18.27 | *p* < .0001 |  |
| GDP *per capita* | -0.0015 | 0.00023 | -6.63 | *p* <.0001 |  |
| Policy 2017 | -1.71 | 1.17 | -1.46 | *p* = .15 |  |
| Policy 2018 | 1.63 | 1.31 | 1.25 | *p* = .21 |  |
| Seasonality (smooth term) |  |  |  | *p* < .001 |  |
| AR(1) MA(0) |  |  |  |  |  |

### Supplementary Table S3: Models of MLDA, taxation, and economic wealth on mortality rates among 18- to 19-year-old men in Lithuania

|  | Estimated effect | Standard Error | t-value | p-value | Adjusted-R^2^ |
| --- | --- | --- | --- | --- | --- |
| Model 1 (Men) |  |  |  |  | 0.11 |
| Intercept | 12.92 | 0.73 | 17.59 | *p* < .0001 |  |
| Policy 2018 | -6.02 | 2.24 | -2.69 | *p* = .0073 |  |
| Seasonality (smooth term) |  |  |  | *p* < .0001 |  |
| AR(2) MA(1) |  |  |  |  |  |

| Model 2 (Men) |  |  |  |  | 0.27 |
| --- | --- | --- | --- | --- | --- |
| Intercept | 19.90 | 1.38 | 14.36 | *p* < .0001 |  |
| GDP | -0.0025 | 0.00045 | -5.64 | *p* <.0001 |  |
| Policy 2017 | -2.34 | 2.29 | -1.02 | *p* = .31 |  |
| Policy 2018 | 1.10 | 2.58 | 0.43 | *p* = .67 |  |
| Seasonality (smooth term) |  |  |  | *p* < .0002 |  |
| AR(2) MA)(1) |  |  |  |  |  |

| Model 3 (Men) |  |  |  |  | 0.27 |
| --- | --- | --- | --- | --- | --- |
| Intercept | 20.17 | 1.38 | 14.66 | *p* < .0001 |  |
| GDP | -0.0025 | 0.00045 | -5.64 | *p* <.0001 |  |
| Policy 2018 | -0.88 | 1.74 | -0.51 | *p* = .61 |  |
| Seasonality (smooth term) |  |  |  | *p* < .0002 |  |
| AR(2) MA)(1) |  |  |  |  |  |

### Supplementary Table S4: Models of MLDA and economic wealth on mortality rates among 15- to 17-year-olds in Lithuania

|  | Estimated effect | Standard Error | t-value | p-value | Adjusted-R^2^ |
| --- | --- | --- | --- | --- | --- |
| Model 1 (Total) |  |  |  |  | 0.11 |
| Intercept | 4.41 | 0.22 | 19.67 | *p* < .0001 |  |
| Policy 2018 | -1.20 | 0.68 | -1.76 | *p* = .079 |  |
| Seasonality (smooth term) |  |  |  | *p* < .0001 |  |
| AR(5) MA(1) |  |  |  |  |  |

| Model 2(Total) |  |  |  |  | 0.22 |
| --- | --- | --- | --- | --- | --- |
| Intercept | 6.24 | 0.46 | 13.60 | *p* < .0001 |  |
| GDP | -0.00061 | 0.00015 | -3.97 | *p* <.0001 |  |
| Policy 2017 | -1.46 | 0.76 | -1.90 | *p* = 0.059 |  |
| Policy 2018 | 1.26 | 0.86 | 1.47 | *p* = .14 |  |
| Seasonality (smooth term) |  |  |  | *p* < .0001 |  |
| AR(5) MA)(1) |  |  |  |  |  |

| Model 3(Total) |  |  |  |  | 0.21 |
| --- | --- | --- | --- | --- | --- |
| Intercept | 6.38 | .48 | 13.35 | *p* < .0001 |  |
| GDP | -0.00068 | 0.00016 | -4.40 | *p* <.0001 |  |
| Policy 2018 | 0.05 | .60 | 0.08 | *p* = .92 |  |
| Seasonality (smooth term) |  |  |  | *p* < .0001 |  |
| AR(5) MA)(1) |  |  |  |  |  |

### Supplementary Table S5: Models of MLDA and economic wealth on mortality rates among 20- to 22-year-olds in Lithuania

|  | Estimated effect | Standard Error | t-value | p-value | Adjusted-R^2^ |
| --- | --- | --- | --- | --- | --- |
| Model 1(Total) |  |  |  |  | 0.17 |
| Intercept | 9.46 | 0.33 | 28.95 | *p* < .0001 |  |
| Policy 2018 | -3.65 | 1.00 | -3.64 | *p* = .00033 |  |
| Seasonality (smooth term) |  |  |  | *p* < .0001 |  |
| AR(0) MA(2) |  |  |  |  |  |

| Model 2(Total) |  |  |  |  | 0.42 |
| --- | --- | --- | --- | --- | --- |
| Intercept | 14.32 | 0.70 | 20.33 | *p* < .0001 |  |
| GDP | -0.0016 | 0.00023 | -7.02 | *p* <.0001 |  |
| Policy 2017 | -2.63 | 1.17 | -2.24 | *p* = .026 |  |
| Policy 2018 | 2.05 | 1.31 | -1.56 | *p* = .12 |  |
| Seasonality (smooth term) |  |  |  | *p* < .0001 |  |
| AR(0) MA(2) |  |  |  |  |  |

| Model 3(Total) |  |  |  |  | 0.41 |
| --- | --- | --- | --- | --- | --- |
| Intercept | 14.62 | 0.71 | 20.60 | *p* < .0001 |  |
| GDP | -0.0018 | 0.00023 | -7.78 | *p* <.0001 |  |
| Policy 2018 | -0.15 | 0.90 | -0.17 | *p* = .87 |  |
| Seasonality (smooth term) |  |  |  | *p* < .0002 |  |
| AR(0) MA(2) |  |  |  |  |  |

### Supplementary Table S6: Estimated trends of mortality rate during four different periods of alcohol control policy in Lithuania among 18- to 19-year-olds

|  | Estimated effect | Standard Error | t-value | p-value | Adjusted-R^2^ |
| --- | --- | --- | --- | --- | --- |
| Model 1  (Total, aged 18 – 19) |  |  |  |  | 0.20 |
| Intercept | 8.95 | 0.52 | 17.08 | *p* < .0001 |  |
| Period 1  Jan 2001 – Dec 2007 | 0.013 | 0.012 | 0.60 | *p* = .30 |  |
| Period 2  Jan 2008 – Dec 2009 | - 0.041 | 0.069 | - 0.60 | *p* = 0.55 |  |
| Period 3  Jan 2010 – Mar 2014 | - 0.062 | 0.024 | - 2.61 | *p =* 0.0098 |  |
| Period 4  Apr 2014 – Dec 2019 | - 0.080 | 0.017 | - 4.80 | *p* < .0001 |  |
| Seasonality (smooth term) |  |  |  | *p* < .033 |  |
| AR(1) MA(0) |  |  |  |  |  |

| Model 2  (Total, aged 18 – 19) |  |  |  |  | 0.16 |
| --- | --- | --- | --- | --- | --- |
| Intercept | 8.46 | 0.55 | 15.34 | *p* < .0001 |  |
| Period 1  Jan 2001 – Dec 2007 | 0.022 | 0.014 | 1.62 | *p* = .11 |  |
| Period 2  Jan 2008 – Dec 2009 | -0.0096 | 0.072 | -0.13 | *p* = 0.90 |  |
| Period 3  Jan 2010 – Feb 2017 | -0.031 | 0.013 | - 2.35 | *p =* 0.020 |  |
| Period 4  Mar 2017 – Dec 2019 | - 0.15 | 0.045 | -3.37 | *p* = 0.00088 |  |
| Seasonality (smooth term) |  |  |  | *p* < .04 |  |
| AR(1) MA(0) |  |  |  |  |  |

### Supplementary Table S7: Slope during four different periods of alcohol control policy in Lithuania among 18- to 19-year-old men

|  | Estimated effect | Standard Error | t-value | p-value | Adjusted-R^2^ |
| --- | --- | --- | --- | --- | --- |
| Model 1  (Men, aged 18 – 19) |  |  |  |  | 0.25 |
| Intercept | 14.13 | 0.99 | 14.26 | *p* < .0001 |  |
| Period 1  Jan 2001 – Dec 2007 | 0.023 | 0.024 | 0.94 | *p* = 0.35 |  |
| Period 2  Jan 2008 – Dec 2009 | -0.093 | 0.13 | -0.72 | *p* = 0.48 |  |
| Period 3  Jan 2010 – Mar 2014 | -0.10 | 0.044 | -2.39 | *p* = 0.018 |  |
| Period 4  Apr 2014 – Dec 2019 | -0.14 | 0.032 | -4.48 | *p* < .0001 |  |
| Seasonality (smooth term) |  |  |  | *p* < .0031 |  |
| AR (2) MA(1) |  |  |  |  |  |

| Model 2  (Men, aged 18 – 19) |  |  |  |  | 0.20 |
| --- | --- | --- | --- | --- | --- |
| Intercept | 13.16 | 1.11 | 11.84 | *p* < .0001 |  |
| Period 1  Jan 2001 – Dec 2007 | 0.038 | 0.027 | 1.38 | 0.17 |  |
| Period 2  Jan 2008 – Dec 2009 | -0.030 | 0.15 | -0.20 | *p* = 0.84 |  |
| Period 3  Jan 2010 – Feb 2017 | -0.044 | 0.026 | -1.71 | *p =* 0.089 |  |
| Period 4  Mar 2017 – Dec 2019 | -0.28 | 0.091 | -3.12 | *p =* 0.0021 |  |
| Seasonality (smooth term) |  |  |  | *p* < .0002 |  |
| AR (2) MA(1) |  |  |  |  |  |
